# The impact of the first UK Covid-19 lockdown on carers and people living with low prevalence dementia: results from the Rare Dementia Support survey

**DOI:** 10.1101/2020.12.18.20248455

**Authors:** Aida Suárez-González, Emma Harding, Nikki Zimmerman, Zoe Hoare, Emilie Brotherhood, Sebastian Crutch

## Abstract

**Introduction:** The public health measures imposed to contain Covid-19 during the first UK lockdown resulted in significant changes in the provision of community support and care for people with dementia. People with low prevalence and young-onset dementias often experience non-memory, behavioural or neuropsychiatric symptoms that require specialised support.

**Objective:** We explored the impact of the first Covid-19 lockdown on people living with low prevalence and young-onset dementia and their carers in the UK.

**Method:** An online survey, including eleven questions about the impact of the lockdown on both the person with dementia and their family caregivers was conducted. Participants were people living with dementia and caregivers who are members of the UK national-reach organisation Rare Dementia Support.

**Results:** 184 carers and 24 people with dementia completed the survey. People with dementia experienced worsening of cognitive symptoms (70%), ability to do things (62%) and well-being (57%) according to their carers. Carers also reported a reduction in the support received for caring (55%). 93% of carers of people living in care homes reported a reduction in their ability to provide care. 26% of carers reported changes in the medication of the person with dementia during the lockdown. 74% of people with dementia reported decreased ability to connect with people socially.

**Conclusions:** People with dementia experienced a worsening of dementia symptoms, removal of support and increased difficulty to connect with other people socially during the 1^st^ wave of Covid-19. Carers encountered barriers to both receiving and providing support and a decline in their own mental health and well-being.

**Key points:**

- 70 % of carers reported **cognitive symptoms getting worse** during the lockdown (e.g., the person with dementia being more disoriented and finding it more difficult to communicate).
- 26 % of carers reported a **change (initiation or increase) in medication** in the person with dementia during the lockdown.
- 79 % carers reported **their own physical or mental health getting worse** due to the lockdown. This increased to 93% when considering responses only from family carers of people living in care homes.
- 93 % of family carers of **people living in care homes** found it harder to continue providing care and support for their relative due to Covid-19.

## 1. Introduction

The United Kingdom declared a nation-wide lockdown due to the Covid-19 pandemic on 23^rd^ March 2020. The measures that followed involved a stay-at-home order, banning of all non-essential contact with people outside of one’s household and suspension of all non-essential services. Easing of restrictions started at the beginning of June 2020, more than two months after the lockdown was declared. The policies to contain Covid-19 impacted many vulnerable groups in society but particularly people living with dementia (Suárez-Gonzalez et al., 2020). For instance, care homes (where 86% of residents have dementia (Livingstone et al., 2017)) experienced high rates of Covid-19 infections and deaths during the pandemic (Bell et la., 2020). 49.5% of those Covid-19 deaths occurred in people living with dementia (ONS, 3 July 2020). Care homes implemented strict infection control guidelines to prevent and contain the spread of infection. These guidelines included a ban on visits which prevented access to family caregivers and led to high levels of isolation in residents. Primary sources of support for people with dementia living in the community were also removed or significantly reduced during the lockdown and the months that followed. The physical distancing and infection control measures imposed changes in routines, raised barriers for people with dementia to access therapies and health care, and for carers to obtain support in caring. The lockdown and the first wave of Covid-19, in general, has had a detrimental impact on people with dementia, both for those living in the community and care homes. The main changes reported in people living with dementia under lockdown are worsening of behavioural and psychological symptoms (including apathy, depression, agitation, anxiety and irritability and walking with purpose), followed by decline in functional abilities for daily living (Lara et al., 2020; Canevelli et al., 2020; Cohen et al., 2020; Boutoleau-Bretonniere et al., 2020; Cagning et al., 2020) and increased used of antipsychotics (Howard et al., 2020). For those in care homes (usually in later stages of the disease and with more severe symptoms) a decline in cognition and ability to perform activities of daily living and an increase in depression and anxiety have been reported (El Haj et al., 2020; O’Caoimh et al., 2020).

Rare Dementia Support (RDS) is an organisation led by the Dementia Research Centre at University College London. It offers education and peer support groups to people with low prevalence and young-onset dementias: posterior cortical atrophy (PCA) (Tang-Wai et al., 2004, Crutch et al., 2017), primary progressive aphasia (PPA) (Gorno-Temppini et al., 2011), behavioural variant Fronto Temporal Dementia (bvFTD) (Racovsky et al., 2011), dementia with Lewy bodies (DLB) (McKeith et al., 2017) and familial Alzheimer’s disease (fAD) (Bateman et al., 2011) and FTD (fFTD) (Greaves & Rohrer, 2019). It also offers a direct support service provided by three highly specialist support workers, and an Admiral Nurse, and has 2000 members, 1850 of whom are people living with dementia and family/friends, while the rest are professionals. People living with these types of dementias face particular challenges during Covid-19 related to their younger age (not fitting within societal perceptions of dementia) and phenotype-specific symptoms (Suárez-González et al., 2020). The clinical presentation of these dementias differs from the canonical episodic memory decline that characterises typical and sporadic forms of Alzheimer’s disease. For instance, PCA is characterised by a progressive deterioration of the visual processing system, resulting in difficulties with reading, reaching for and locating objects and issues with spatial navigation. People living with PPA experience a progressive deterioration in language and communication, with language difficulties the dominant symptom during the early stages of the disease. People with bvFTD show changes in social behaviour, judgment, impulsivity and empathy. The progressive cognitive decline of people with dementia with Lewy bodies is accompanied by visual hallucinations, fluctuations and/or parkinsonism. Lastly, familial forms of both AD and FTD are caused by autosomal dominant genetic mutations (meaning there is a 50% chance a child of a carrier parent will inherit it) transmitted through families. Carriers of these mutations show symptoms at a very young age, often in their 40s or 50s.

People with these dementias and their families require specialised care and support (Millenaar et al., 2016) and they may also experience particular challenges during the pandemic. The goal of this study was to describe the lockdown’s impact among members of RDS who live with low prevalence and young-onset dementia and their caregivers.

## 2. Methods

The reporting of the method and results from this survey complies with the CHERRIES survey reporting checklist (Eysenbach, 2004). This study was conducted as part of the RDS Impact study (Brotherhood et al., 2020) and granted permission by the UCL Research Ethics Committee (8545/004: RDS Impact Study).

### 2.1. Survey development

We built an online survey made of 11 questions (one per page) about the impact of the lockdown on the person with dementia’s cognitive symptoms, psychological well-being, ability to do things, ability to connect with people socially, general health and changes in medication (see Appendix 1). It also included questions about the health of the caregiver and support received in caring, helpful strategies developed during the lockdown and any positives found in the situation. The first eight questions of the survey could be responded to by selecting “yes”, “no” or “not sure” (e.g., *Have the person’s cognitive symptoms got worse during the lockdown (for example being more disoriented, finding it more difficult to communicate)*. If respondents marked ‘yes’ in one of the first eight questions, a comment box was provided for the participant to give further details (e.g., *If yes, please tell us how*). The last three questions were open, for the respondent to elaborate about a) helpful strategies to cope with lockdown, b) positive aspects of the lockdown (if any) and c) any other additional information about their experience during the lockdown that the respondent wished to share.

The questions were informed by emerging evidence from research in the field (Lara et al., 2020; Canevelli et al., 2020; Cohen et al., 2020; Boutoleau-Bretonniere et al., 2020; Cagning et al., 2020) and feedback from the experience of the Direct Support Team at RDS and clinical experience. Three variations in the survey questions were produced to suit the three types of respondents approached:

1. People living with dementia
2. Carers of people with dementia living in the community and
3. Carers of people living in care homes.

The electronic questionnaire was built in Qualtrics software version July-August 2020 (Qualtrics, Provo, UT) and tested for technical functionality and usability by three members of the research team (ASG, NZ, EH).

### 2.2. Recruitment and procedure

The survey was distributed to the RDS mailing list on 11^th^ of August and remained open until 30^th^ of September. Participants were members of RDS who were living with or caring for someone with low prevalence dementia. RDS members received an email explaining the purpose of the survey (wording of this email is available in Appendix 2) and providing instructions for completion along with a link to access the survey. Participants were provided a link on the survey homepage to the Participant Information Sheet, and were required to opt in to the survey confirming this had been read, understood, and the participant was happy to proceed and provide responses. Any member registered in the RDS mailing database who was either a person living with dementia or a carer was given the opportunity to participate. This was an open survey since no login was needed to access the survey. However, only RDS members had access to it.

No personal information was collected in this survey, and no incentives were offered for survey completion. A response was not required to proceed to the next page(respondents could progress onwards leaving some questions blank) and respondents were able to go back to check and edit previous entries. Questions were not randomised. The survey was set up so that each respondent could only use the survey link once, and each completed questionnaire was allocated a unique ID. Respondents had to proceed to the end of the survey for their response to be submitted. Qualtrics allowed automatic capture of responses.

### 2.3. Analysis

The current study presents the quantitative data from the first eight questions. Resultant data are being presented descriptively and not statistically tested as the survey was designed to be exploratory of the population rather than hypothesis driven. Responses are expressed in frequency counts (percentages) and contingency tables. Data are classified according to respondent group (carer or person living with dementia), subgroup (living in the community together, living in the community but not together and living in a care home) and phenotype. A qualitative analysis of responses to the survey’s open questions is underway for write up in an independent paper (Harding et al. 2020, in preparation).

## 3. Results

An email with survey details was sent to 1850 members including people living with dementia and carers. We obtained 208 completed surveys. 184/208 of respondent were caregivers (Table 1) and 24/208 people living with dementia (Table 2).

**Table 1.** Responses of carers to the Covid-19 lockdown survey questions

|  | **Total**<br>**(N= 184)** | **FTD**<br>**(n=62)** | **PCA**<br>**(n=59)** | **PPA**<br>**(n=36)** | **DLB**<br>**(n=10)** | **fFTD**<br>**(n=8)** | **fAD**<br>**(n=3)** | **Other**<br>**(n=6)** |
| --- | --- | --- | --- | --- | --- | --- | --- | --- |
| **Questions about the person with dementia** |  |  |  |  |  |  |  |  |
| Decline in cognitive symptoms | 70% | 66% | 69% | 72% | 80% | 75% | 100% | 67% |
| Negative impact on behaviour or wellbeing | 57% | 47% | 64% | 53% | 70% | 63% | 100% | 83% |
| Loss ability to do things | 62% | 55% | 66% | 50% | 90% | 88% | 67% | 83% |
| General health affected | 47% | 38% | 45% | 53% | 70% | 71% | 100% | 67% |
| Medication changes† | 26% | 23% | 29% | 18% | 50% | 25% | 0% | 50% |
| Ability to connect with people socially | 71% | 60% | 78% | 75% | 90% | 75% | 67% | 67% |
| **Questions about the carer** |  |  |  |  |  |  |  |  |
| Level of stress or health impacted by the lockdown | 79% | 72% | 80% | 83% | 90% | 75% | 100% | 83% |
| Negative impact on support received for caring | 55% * | 52% | 58% | 58% | 50% | 40% | 100% | 50% |
| Harder for carer to provide care and support | 93% ** | 92% | 93% | 67% | 100% | 100% | 100% | 100% |

\*N=144, including only carers of people living in the community
\*\*N=40, including only carers of people living in care homes
† increase or initiation of antidepressants, benzodiazepines, antipsychotics, “sleep medication” (as reported by respondents) and pain killers and in less frequently reported blood thinners, anticholinesterase inhibitors and beta blockers.
Red cells = equal or more than 75% of “yes” responses
Yellow cells = between 50 – 74% of “yes” responses

**Table 2.** Responses of people living with dementia

|  | **Total**<br>**(N= 24)** | **FTD**<br>**(n=5)** | **PCA**<br>**(n=8)** | **PPA**<br>**(n=7)** | **DLB**<br>**(n=3)** | **fFTD**<br>**(n=0)** | **fAD**<br>**(n=1)** | **Other**<br>**(n=0)** |
| --- | --- | --- | --- | --- | --- | --- | --- | --- |
| Decline in cognitive symptoms | 38% | 40% | 38% | 43% | 33% | - | 0% | - |
| Negative impact on behaviour or wellbeing | 50% | 80% | 38% | 29% | 100% | - | 0% | - |
| Loss ability to do things | 46% | 60% | 38% | 43% | 67% | - | 0% | - |
| General health affected | 42% | 40% | 38% | 29% | 100% | - | 0% | - |
| Medication changes | 13% | 0% | 0% | 14% | 33% | - | 100% | - |
| Support received | 54% | 40% | 63% | 57% | 33% | - | 100% | - |
| Ability to connect with people socially | 74% | 40% | 88% | 83% | 67% | - | 100% | - |

Red cells = equal or more than 75% of “yes” responses
Yellow cells = between 50 – 74% of “yes” responses

Overall, 70% of the carers reported a decline in cognitive symptoms and ability to connect with people socially and a negative impact on their level of stress and health during the first lockdown in the UK. Medication changes were reported in 26% of the cases. This involved increase or initiation of antidepressants, benzodiazepines, antipsychotics, “sleep medication” (as reported by respondents) and pain killers and in less frequently reported blood thinners, anticholinesterase inhibitors and beta blockers

74% of people with dementia surveyed reported increased difficulties in connecting with people socially due to the lockdown and 50% said that the lockdown negatively impacted their well-being and the support received.

### Carers of people in the community living together

Table 3 summarises responses of 120/184 carers who were living together with the person with dementia when completing the survey. 40/120 were carers of people living with PCA, while 38 and 25 cared for people with FTD and PPA respectively. A minority of respondents were carers of people with DLB, fFTD, fAD and people with other diagnoses (e.g., corticobasal syndrome or respondents who were unsure about diagnosis. Across all phenotypes, at least 70% of carers reported decline in the person with dementia’s cognitive symptoms and ability to connect with other people socially during the lockdown, and a negative impact on their own health or stress level. The most frequently reported change among carers of people with FTD and PPA was a worsening of carers’ health and level of stress (64% and 76% respectively). For carers of people with PCA it was the loss of the ability of the person with dementia to connect socially (80%). 33% of carers of people with DLB reported changes in medication during the lockdown.

**Table 3.** Responses of carers living together in the community with the person with dementia.

|  | **Total**<br>**(N= 120)** | **FTD**<br>**(n=38)** | **PCA**<br>**(n=40)** | **PPA**<br>**(n=25)** | **DLB**<br>**(n=6)** | **fFTD**<br>**(n=5)** | **fAD**<br>**(n=2)** | **Other**<br>**(n=4)** |
| --- | --- | --- | --- | --- | --- | --- | --- | --- |
| **Questions about the person with dementia** |  |  |  |  |  |  |  |  |
| Decline in cognitive symptoms | 70% | 61% | 78% | 68% | 83% | 80% | 100% | 50% |
| Negative impact on behaviour or wellbeing | 58% | 50% | 73% | 44% | 67% | 80% | 0% | 75% |
| Loss ability to do things | 61% | 50% | 73% | 44% | 100% | 80% | 50% | 75% |
| General health affected | 44% | 38% | 43% | 44% | 67% | 80% | 0% | 50% |
| Medication changes | 24% | 22% | 28% | 16% | 33% | 20% | 0% | 50% |
| Ability to connect with people socially | 72% | 64% | 80% | 68% | 100% | 80% | 50% | 50% |
| **Questions about the carer** |  |  |  |  |  |  |  |  |
| Level of stress or health impacted by the lockdown | 73% | 68% | 73% | 76% | 83% | 80% | 100% | 75% |
| Negative impact on support received for caring | 59% | 54% | 65% | 60% | 50% | 40% | 100% | 50% |

Red cells = equal or more than 75% of “yes” responses
Yellow cells = between 50 – 74% of “yes” responses

### Carers of people in the community not living together

Table 4 summarises the responses of 24/184 carers of people with dementia who did not live together in the same household at the time of survey completion. 11/24 were carers of people with FTD, 8 PPA and 5 PCA. More than 80% of the carers reported loss of the ability of the person with dementia to connect with people socially and a worsening of their own stress levels or health. The most frequently reported impact in the FTD group was the person with dementia losing the ability to do things and their ability to connect socially. This was similar in the PCA group, in addition to 5/5 carers reporting worsening in their own stress/health. All carers of people with PPA reported a loss of ability to connect with other people socially and increased levels of stress/deterioration of caregiver health. 75% of PPA caregivers also reported a further decline in cognitive symptoms, well-being or behaviour and general health of the person with dementia.

**Table 4.** Responses of carers of people with dementia living in the community but not together

|  | **Total**<br>**(N= 24)** | **FTD**<br>**(n=11)** | **PCA**<br>**(n=5)** | **PPA**<br>**(n=8)** | **DLB**<br>**(n=0)** | **fFTD**<br>**(n=0)** | **fAD**<br>**(n=0)** | **Other**<br>**(n=)** |
| --- | --- | --- | --- | --- | --- | --- | --- | --- |
| **Questions about the person with dementia** |  |  |  |  |  |  |  |  |
| Decline in cognitive symptoms | 63% | 64% | 40% | 75% | - | - | - | - |
| Negative impact on behaviour or wellbeing | 63% | 64% | 40% | 75% | - | - | - | - |
| Loss ability to do things | 71% | 73% | 80% | 63% | - | - | - | - |
| General health affected | 52% | 36% | 50% | 75% | - | - | - | - |
| Medication changes | 29% | 36% | 20% | 25% | - | - | - | - |
| Ability to connect socially | 88% | 73% | 100% | 100% |  |  |  |  |
| **Questions about the carer** |  |  |  |  |  |  |  |  |
| Level of stress or health impacted by the lockdown | 83% | 64% | 100% | 100% | - | - | - | - |
| Negative impact on support received for caring | 38% | 45% | 0% | 50% | - | - | - | - |

Red cells = equal or more than 75% of “yes” responses
Yellow cells = between 50 – 74% of “yes” responses

### Carers of people living in care homes

Table 5 summarises the responses of 40/184 carers whose relative with dementia was living in a care home at the time of responding to the survey. 93% of carers reported that the lockdown increased their level of stress and that it became more challenging for them to provide care and support to their relative living in the care home. The majority of respondents were carers of people with FTD and PCA. The most commonly reported change in these two groups was a decline in cognition (85%) and the ability to connect with people socially (64%) respectively.

**Table 5.** Responses of carers of people with dementia living in care homes

|  | **Total**<br>**(N= 40)** | **FTD**<br>**(n=13)** | **PCA**<br>**(n=14)** | **PPA**<br>**(n=3)** | **DLB**<br>**(n=4)** | **fFTD**<br>**(n=3)** | **fAD**<br>**(n=1)** | **Other**<br>**(n=2)** |
| --- | --- | --- | --- | --- | --- | --- | --- | --- |
| **Questions about the person with dementia** |  |  |  |  |  |  |  |  |
| Decline in cognitive symptoms | 75% | 85% | 57% | 57% | 75% | 67% | 100% | 100% |
| Negative impact on behaviour or wellbeing | 48% | 23% | 50% | 50% | 75% | 33% | 100% | 100% |
| Loss ability to do things | 60% | 54% | 43% | 43% | 75% | 100% | 100% | 100% |
| General health affected | 54% | 38% | 50% | 50% | 75% | 50% | 100% | 100% |
| Medication changes | 30% | 15% | 36% | 36% | 75% | 33% | 0% | 50% |
| Ability to connect socially | 60% | 38% | 64% | 64% | 75% | 67% | 100% | 100% |
| **Questions about the carer** |  |  |  |  |  |  |  |  |
| Level of stress or health impacted by the lockdown | 93% | 92% | 93% | 93% | 100% | 67% | 100% | 100% |
| Harder for carer to provide care and support | 93% | 92% | 93% | 93% | 100% | 100% | 100% | 100% |

Red cells = equal or more than 75% of “yes” responses
Yellow cells = between 50 – 74% of “yes” responses

## 4. Discussion

This is the most extensive survey documenting the impact of the first nationwide UK Covid-19 lockdown on people living with low prevalence and young-onset dementias. The majority of carers surveyed reported worsening of health and cognitive symptoms of the person with dementia due to the Covid-19 restrictions. Almost all caregivers of people living in care homes said it was harder for them to provide support under current circumstances. More than half of carers also reported that the person with dementia’s ability to connect socially diminished, along with the ability to do things, and a worsening of well-being and behavioural symptoms. The majority of people with dementia expressed a loss in their ability to connect socially and around half also expressed a decline in the support received during the lockdown and a detrimental effect on their well-being.

### Carers of people living in the community

The majority of respondents were carers of people with FTD, PCA and PPA. Differences in the number of respondents by diagnostic group reflect the composition of membership in RDS. Our results are in keeping with recent studies describing the impact of lockdown on people living with FTD and DLB. Cagnin et al. (Cagnin et al., 2020) surveyed 775 carers from 87 Italian memory clinics. They found that 62% of people with DLB and 55% with FTD experienced the onset or worsening of psychological and behavioural symptoms.

Carers of people in the community living together made the largest group of respondents, probably because it is also the most represented group of RDS members. We observed that carers of people living in care homes reported more changes than those living in the community separately, who in turn reported more changes than carers living in the community together with the person with dementia. This observation may be driven by sample size but also it may be partially explained by several reasons. On the one hand, people with dementia that live away from their carer relative may have received less support during the lockdown, which may have accelerated decline. It might also be that carers who cannot see their relatives experience more anxiety and a higher subjective perception of decline. Additionally, carers who live with their relatives with dementia might report fewer changes because they see the person every day and subtle differences go unnoticed, while those who were seeing the person less often may have noticed changes more easily.

### Carers of people living in care homes

A survey about the restriction of visitors in Irish care homes revealed that 54% of people with dementia experienced a worsening in their memory and 43% a decline in function (O’Caoimh et al., 2020). In our sample, the worsening of cognitive symptoms reaches 75% and decline in function 60%. Differences may be attributable to the use of different methods and tools and lack of comparability of samples.

The prolonged separation resulting from the ban on visits during lockdown left carers of people living in care homes unable to provide care to their relatives and this might be taking a toll on their mental health. Moreover, this group also represents the higher proportion of respondents reporting worsening of cognitive symptoms in the person with dementia, in particular for those living with DLB.

### Medication

Changes in medication were reported in 26% of the cases. This is close to the 27% increase reported by Cagnin et al., (Cagnin et al., 2020), the 20% increase reported by Cohen et al. (Cohen et al., 2020) and far higher than the 7% reported by Canevelli et al. (Cavenelli et al., 2020). As in previous studies, many of the changes had to do with commencement or increase in the use of antipsychotics, benzodiazepines and antidepressants. This indicates a worsening in mental health and psychological symptoms and resonates with the responses to those questions in the survey.

### Limitations

This study documents the experience of members of RDS, a highly specialised organisation providing support to people affected by low prevalence and young-onset dementias. Some methodological limitations should be acknowledged. First, no demographics were collected in this survey, which impedes comparisons across groups (since results could not be interpreted nor confounding variables considered). Second, limitations also apply to the representativeness of this sample and drawing generalisable conclusions. For instance, it may be that people who felt more stressed were also more motivated to share their experience and therefore, may be overrepresented. The opposite is also possible that people who felt more stressed were less available and therefore are underrepresented. It may also be that people who are more engaged with the activities of RDS also feel closer to the staff and researchers, and this familiarity may have prompted more particiaption among those members. Third, only people able to use technology could engage in the online survey. This is an important limitation for many, with deeper repercussions in the representativeness of data collected online but also equity and equality in accessing research and other services. Fourth, people with dementia were also encouraged to participate in the survey. However, no system was in place to support their participation, which biases both the kind of people with dementia who could respond to the survey and the responses themselves. For instance, only 38% of people with dementia reported changes in cognitive status, while 70% of carers did. This may be because only people in mild stages of the disease were able to engage in the survey or because of poor awareness of the cognitive changes experienced. Barriers to engaging and participating digitally remain a big challenge. Finally, the questions in the survey were framed negatively, which may have favoured negative responses. However, an additional open question for people to report positives was also included.

## 5. Conclusions

This paper intends to document the impact of the first UK Covid-19 lockdown on people living with low prevalence and young-onset dementias so that their needs are considered within national responses in future waves. The Covid-19 pandemic has negatively impacted the health and well-being of the majority of carers and people with low prevalence and young-onset dementia who participated in this survey. Social and cognitive stimulation and specialised therapeutic support are essential to enable people to live well with dementia, yet these fundamental pillars of the dementia care pathway were interrupted during Covid-19 first wave’s lockdown.

## Data Availability

The data that support the findings of this study are available on request from the corresponding author, [ASG]. The data are not publicly available in compliance with ethic and funding requirements but will be uploaded to a data repository for researchers from different institutions to access after the study ends in 2024.

## References

Bateman, R. J., Aisen, P. S., De Strooper, B., Fox, N. C., Lemere, C. A., Ringman, J. M., & Xiong, C. (2011). Autosomal-dominant Alzheimer’s disease: a review and proposal for the prevention of Alzheimer’s disease. Alzheimer’s research & therapy, 3(1), 1–13.

Bell D, Comas-Herrera A, Henderson D, Jones S, Lemmon E, Moro M, Murphy S, O’Reilly D and Patrignani P (2020) COVID-19 mortality and long-term care: a UK comparison. Article in LTCcovid.org, International LongTerm Care Policy Network, CPEC-LSE, August 2020.

Brotherhood EV, Stott J, Windle G, Barker S, Culley S, Harding E, Camic PM, Caufield M, Ezeofor V, Hoare Z, McKee-Jackson R, Roberts J, Sharp R, Suarez-Gonzalez A, Sullivan MP, Tudor Edwards R, Walton J, Waddington C, Winrow E, Crutch SJ. Protocol for the Rare Dementia Support Impact study: RDS Impact. Int J Geriatr Psychiatry. 2020 Aug;35(8):833–841. doi: 10.1002/gps.5253. Epub 2020 Feb 17. PMID 31876030.

Cagnin A, Di Lorenzo R, Marra C, Bonanni L, Cupidi C, Laganà V, Rubino E, Vacca A, Provero P, Isella V, Vanacore N, Agosta F, Appollonio I, Caffarra P, Pettenuzzo I, Sambati R, Quaranta D, Guglielmi V, Logroscino G, Filippi M, Tedeschi G, Ferrarese C, Rainero I, Bruni AC; SINdem COVID-19 Study Group. Behavioral and Psychological Effects of Coronavirus Disease-19 Quarantine in Patients With Dementia. Front Psychiatry. 2020 Sep 9;11:578015. doi: 10.3389/fpsyt.2020.578015. PMID: 33033486; PMCID: PMC7509598.

Canevelli M, Valletta M, Toccaceli Blasi M, Remoli G, Sarti G, Nuti F, Sciancalepore F, Ruberti E, Cesari M, Bruno G. Facing Dementia During the COVID-19 Outbreak. J Am Geriatr Soc. 2020 Aug;68(8):1673–1676. doi: 10.1111/jgs.16644. Epub 2020 Jun 9. PMID: 32516441; PMCID: PMC7300919.

Cohen G, Russo MJ, Campos JA, Allegri RF. COVID-19 Epidemic in Argentina: Worsening of Behavioral Symptoms in Elderly Subjects With Dementia Living in the Community. Front Psychiatry. 2020 Aug 28;11:866. doi: 10.3389/fpsyt.2020.00866. PMID: 33005158; PMCID: PMC7485090.

Crutch SJ, Schott JM, Rabinovici GD, Murray M, Snowden JS, van der Flier WM, Dickerson BC, Vandenberghe R, Ahmed S, Bak TH, Boeve BF, Butler C, Cappa SF, Ceccaldi M, de Souza LC, Dubois B, Felician O, Galasko D, Graff-Radford J, Graff-Radford NR, Hof PR, Krolak-Salmon P, Lehmann M, Magnin E, Mendez MF, Nestor PJ, Onyike CU, Pelak VS, Pijnenburg Y, Primativo S, Rossor MN, Ryan NS, Scheltens P, Shakespeare TJ, Suárez González A, Tang-Wai DF, Yong KXX, Carrillo M, Fox NC; Alzheimer’s Association ISTAART Atypical Alzheimer’s Disease and Associated Syndromes Professional Interest Area. Consensus classification of posterior cortical atrophy. Alzheimer’s & Dementia. 2017 Aug;13(8):870–884. doi: 10.1016/j.jalz.2017.01.014.

O’Caoimh R, O’Donovan MR, Monahan MP, Dalton O’Connor C, Buckley C, Kilty C, Fitzgerald S, Hartigan I, Cornally N. Psychosocial Impact of COVID-19 Nursing Home Restrictions on Visitors of Residents With Cognitive Impairment: A Cross-Sectional Study as Part of the Engaging Remotely in Care (ERiC) Project. Front Psychiatry. 2020 Oct 26;11:585373. doi: 10.3389/fpsyt.2020.585373. PMID: 33192731; PMCID: PMC7649131.

Eysenbach G. Improving the quality of Web surveys: the Checklist for Reporting Results of Internet E-Surveys (CHERRIES) [published correction appears in doi:10.2196/jmir.2042]. J Med Internet Res. 2004;6(3):e34. Published 2004 Sep 29. doi:10.2196/jmir.6.3.e34

Gorno-Tempini, M. L., Hillis, A. E., Weintraub, S., Kertesz, A., Mendez, M., Cappa, S. F., Ogar, J. M., Rohrer, J. D., Black, S., Boeve, B. F., Manes, F., Dronkers, N. F., Vandenberghe, R., Rascovsky, K., Patterson, K., Miller, B. L., Knopman, D. S., Hodges, J. R., Mesulam, M. M., & Grossman, M. (2011). Classification of primary progressive aphasia and its variants. Neurology, 76(11), 1006–1014. https://doi.org/10.1212/WNL.0b013e31821103e6

Greaves CV, Rohrer JD. An update on genetic frontotemporal dementia. J Neurol. 2019 Aug;266(8):2075–2086. doi: 10.1007/s00415-019-09363-4. Epub 2019 May 22. PMID: 31119452; PMCID: PMC6647117.

Howard R, Burns A, Schneider L. Antipsychotic prescribing to people with dementia during COVID-19. Lancet Neurol. 2020;19(11):892. doi:10.1016/S1474-4422(20)30370-7.

Livingston G, Barber J, Marston L, Rapaport P, Livingston D, Cousins S, Robertson S, La Frenais F, Cooper C. Prevalence of and associations with agitation in residents with dementia living in care homes: MARQUE cross-sectional study. BJPsych Open. 2017 Jul 27;3(4):171–178. doi: 10.1192/bjpo.bp.117.005181. PMID: 28794896; PMCID: PMC5530006.

McKeith, I. G., Boeve, B. F., Dickson, D. W., Halliday, G., Taylor, J. P., Weintraub, D., & Bayston, A. (2017). Diagnosis and management of dementia with Lewy bodies: Fourth consensus report of the DLB Consortium. Neurology, 89(1), 88–100.

Millenaar, J. K., Bakker, C., Koopmans, R. T., Verhey, F. R., Kurz, A., & de Vugt, M. E. (2016). The care needs and experiences with the use of services of people with young-onset dementia and their caregivers: a systematic review. International journal of geriatric psychiatry, 31(12), 1261–1276.

ONS (3 July 2020). Deaths involving Covid-19 in the care sector, England and Wales: deaths occurring up to 12 June 2020 and registered up to 20 June 2020. https://www.ons.gov.uk/peoplepopulationandcommunity/birthsdeathsandmarriages/deaths/articles/deathsinvolvingcovid19inthecaresectorenglandandwales/deathsoccurringupto12june2020andregisteredupto20june2020provisional#deaths-involving-covid-19-among-care-home-residents

Rascovsky K, Hodges JR, Knopman D, Mendez MF, Kramer JH, Neuhaus J, Miller BL, et al. Sensitivity of revised diagnostic criteria for the behavioural variant of frontotemporal dementia. Brain. 2011;134:2456–2477.

Suárez-González A, Zimmermann N, Waddington C, Wood O, Harding E, Brotherhood E, Fox NC, Crutch SJ. Non-memory led dementias: care in the time of covid-19. BMJ. 2020 Jun 30;369:m2489. doi: 10.1136/bmj.m2489. PMID: 32606068.

Tang-Wai DF, Graff-Radford NR, Boeve BF, Dickson DW, Parisi JE, Crook R, Caselli RJ, Knopman DS, Petersen RC. Clinical, genetic, and neuropathologic characteristics of posterior cortical atrophy. Neurology. 2004 Oct 12;63(7):1168–74. doi: 10.1212/01.wnl.0000140289.18472.15. PMID: 15477533.

